# National Trends and Resource Utilization Associated with Penetrating Trauma Recidivism

**DOI:** 10.1101/2023.01.09.23284357

**Authors:** Nam Yong Cho, Russyan Mark Mabeza, Syed Shahyan Bakhtiyar, Shannon Richardson, Konmal Ali, Zachary Tran, Peyman Benharash

## Abstract

**BACKGROUND:** While recurrent penetrating trauma has been associated with long-term mortality and disability, national data on factors associated with reinjury remain limited. We examined temporal trends, patient characteristics, and resource utilization associated with repeat firearm-related or stab injury across the US.

**METHODS:** The 2010-2019 Nationwide Readmissions Database was queried to identify all hospitalizations for penetrating trauma. Recidivism was defined as those returned for subsequent penetrating injury within 60 days. We quantified injury severity using the International Classification of Diseases Trauma Mortality Prediction model. Trends in recidivism, length of stay (LOS), hospitalization costs, and rate of non-home discharge were then analyzed. Multivariable regression models were developed to assess the association of recidivism with outcomes of interest.

**RESULTS:** Of an estimated 968,717 patients (28.4% Gunshot, 71.6% Stab), 2.1% experienced recidivism within 60 days of initial injury. From 2010 to 2019, trauma recidivism of gunshot wound increased in annual incidence while that of stab remained stable. Patients experiencing penetrating trauma recidivism were more commonly male, younger and insured by Medicaid. Patients with recidivism had shorter index LOS, lower median index hospitalization cost and higher rates of non-home discharge. After risk adjustment, penetrating trauma recidivism was associated with significantly higher hospitalization costs, shorter time before readmission, and increased odds of non-home discharge. In addition, comorbid conditions of hypothyroidism and psychoses were associated with greater likelihood of stab wound recidivism.

**CONCLUSION:** The trend in penetrating trauma recidivism has been on the rise for the past decade. National efforts to improve post-discharge prevention and social support services for patients with penetrating trauma are warranted and may reduce the burden of recidivism.

## 1. INTRODUCTION

Urban trauma accounts for an estimated 1.4 million annual emergency department visits and nearly $8.5 billion in healthcare expenditures.^1^ Approximately 44% of these admissions are for recurrent injuries in patients with prior injury-related hospitalization, defined as trauma recidivism. Primarily consisting of gunshot and stab injuries, recidivism in penetrating trauma has been linked to increased long-term mortality, with 20% of patients facing death within five years of repeat injury.^2^ Furthermore, individual recidivists with penetrating injuries are likely to present with a second unrelated trauma or death. Such patterns present a critical public health challenge, indicating a need for a thorough analysis of factors associated with trauma recidivism.

Previous institutional studies have noted younger age, Black race, and low-income status to be risk factors for trauma recidivism.^3^ Furthermore, individuals suffering from recurrent penetrating trauma frequently suffer from high morbidity and costs of care due to often required operative interventions and extended inpatient stays. Despite being a significant issue, examination of recidivism in penetrating trauma remains limited to single-center series. Thus, the present study examined national trends as well as clinical and financial outcomes of recidivism in penetrating trauma using a nationally representative cohort. We hypothesized that penetrating recidivism would be associated with psychiatric illnesses and non-home discharge disposition on index hospitalization.

## 2. MATERIAL AND METHODS

### 2.1 Data Source and Study Cohort

This was a retrospective study using 2010-2019 Nationwide Readmissions Database (NRD). Maintained by the Health Care Costs and Utilization Project (HCUP), the NRD is the largest national readmission database in the US and provides accurate national estimates for nearly 60% of all annual hospitalizations. The NRD contains unique identifiers for the patient and hospital variables, which allows readmissions to be tracked within each calendar year.

All adult (≥18 years) hospitalizations for firearm- and stab-related trauma were identified using the *International Classification of Diseases, Ninth/Tenth Revisions* (ICD-9/10) codes (Supplemental Table S1). Entries missing key information, such as sex, age or mortality, were excluded from the analysis (7.5%). We quantified injury severity using the *International Classification of Diseases* Trauma Mortality Prediction Model.^4^ The cohort was stratified by gunshot and stab injuries. In each group, patients were further classified gunshot (*GSW-R*) and stab (*Stab-R*) injury recidivism (Figure 1).

**Figure 1.**
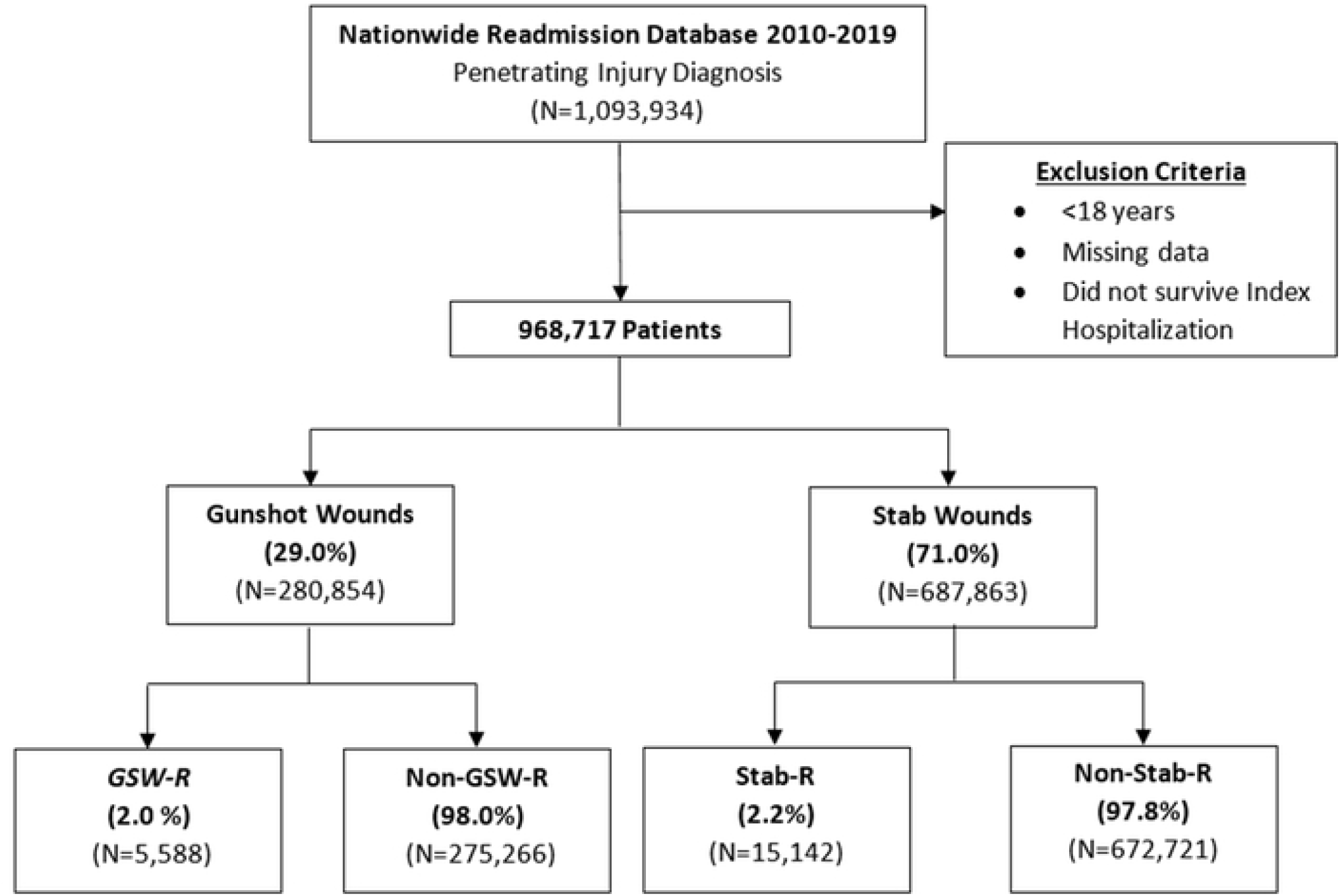
CONSORT (Consolidated Standards of Reporting Trials) diagram of study cohort and survey-weighted sample size. GSW-R, gunshot recidivism. Stab-R, Stab recidivism.

Recidivism was defined as subsequent penetrating injury within 60 days of discharge from the index hospitalization. The timing of 60 days was used to capture at least the 75th percentile of patients experiencing recidivism while minimizing the skewed distribution (Supplemental Figure 1).

### 2.2 Study Variables and Outcomes

Patient and hospital characteristics such as age, insurance status, intent of the injury, hospital region, and income quartiles were defined according to the NRD data dictionary.^5^ The Elixhauser Comorbidity Index, a validated composite score of 30 chronic comorbidities, was used to estimate the burden of comorbidities in the study population.^6^ In addition, specific patient comorbidities were further ascertained using ICD-9/10 codes, including congestive heart failure, diabetes and hypertension, among others (Supplemental Table S1). Mortality analysis was limited to death during hospitalization, whereas non-home discharge was defined as transfer to a post-acute care facility. Hospitalization costs were calculated by applying center-specific cost-to-charge ratios to total hospitalization charges and adjusted for inflation using the 2019 Bureau of Labor Statistics Personal Health Care Price Index. The primary outcome of interest was the trends in penetrating trauma recidivism, while secondary outcomes included index hospitalization length of stay (LOS), cumulative hospitalization costs and non-home discharge.

### 2.3 Statistical Methods

Categorical variables are reported as percentages, and continuous variables as medians with interquartile range (IQR). The Chi-square and Kruskal-Wallis tests were used to compare patient characteristics in both study cohorts. Temporal trends were assessed using a nonparametric test (nptrend). Multivariable regressions were used to identify patient and hospital factors associated with recidivism as well as secondary outcomes. We used Elastic Net regularization to guide variable selection to improve out-of-sample generalizability.^7^ Regression outputs are reported as adjusted odds ratios (AOR) or beta coefficients (β) with 95% confidence intervals (95% CI). A P-value <0.05 was considered statistically significant. All statistical analyses were performed using Stata 16.1 (StataCorp, College Station, TX).

### 2.4 Ethics Statement

As this is a retrospective study of a fully de-identified database, the study did not require informed consent and was deemed exempt from the Institutional Review Board at the University of California, Los Angeles.

## 3. RESULTS

### 3.1 Demographics of Patients and Trends of GSW Recidivism

Of an estimated 280,854 patients, 5,588 (2.0%) experienced recidivism within 60 days of the initial injury. From 2010 to 2019, trauma recidivism of GSW increased in annual incidence (nptrend<0.001, Figure 2). Additionally, non-home discharge among *GSW-R* increased from 16.8% to 20.2% over the study period (nptrend<0.001, Figure 3). Compared to those without recurrent injury, patients in *GSW-R* cohort were more commonly male (88.9 vs 87.0%, P<0.001), younger (30 [23-40] vs 32 [24-44] years, P<0.001), and insured by Medicaid (36.3 vs 32.1%, P<0.001). In addition, patients with recidivism more commonly experienced gunshot injury due to accident (37.2 vs 32.7%, P<0.001) and shorter median time until readmission (12 [4-32] vs 28 [8-87] days, P<0.001), compared to others. Elixhauser comorbidity Index score and prevalence of psychiatric disorders were similar between those with recidivism and others (Table 1).

**Table 1.**
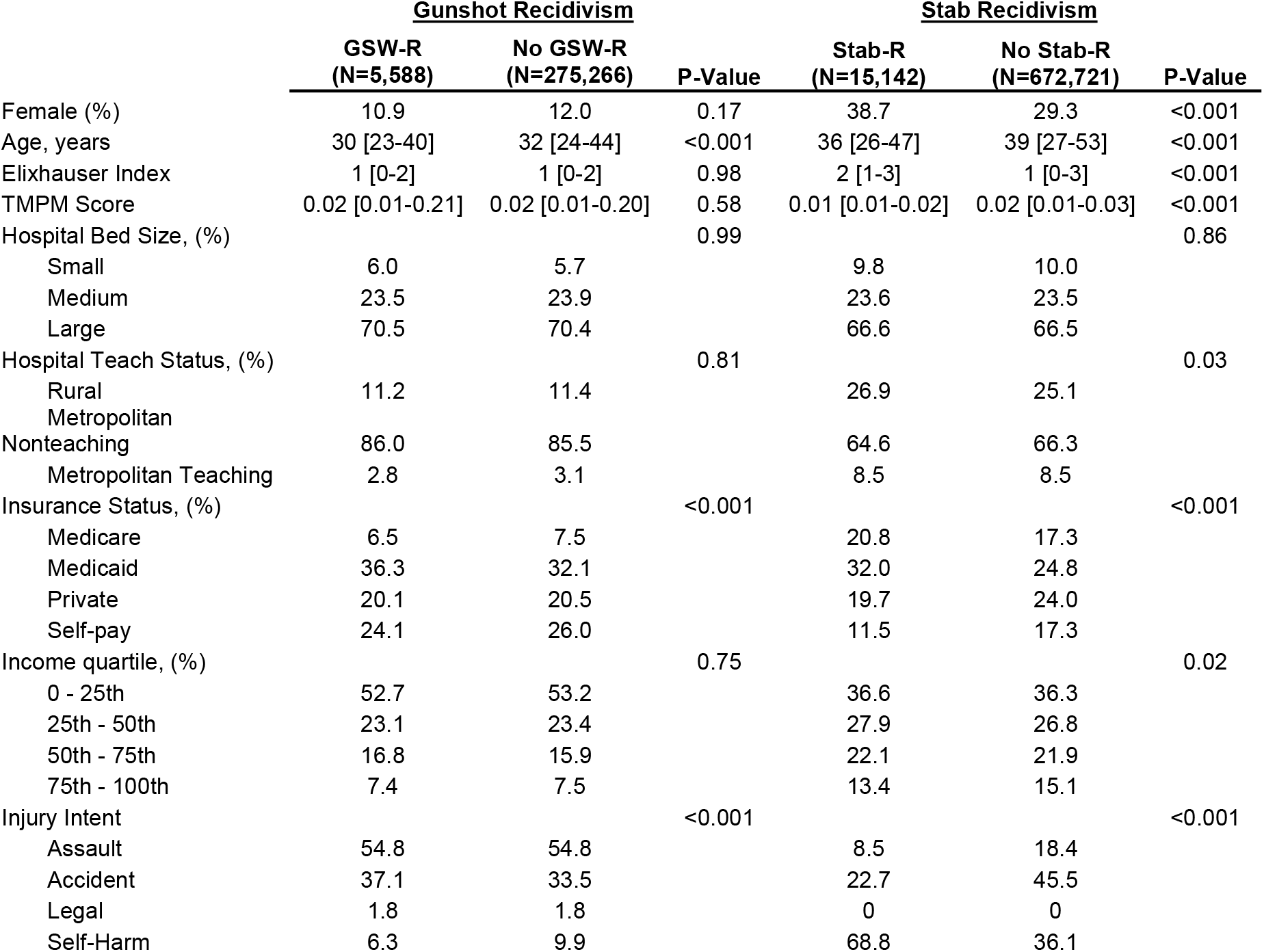
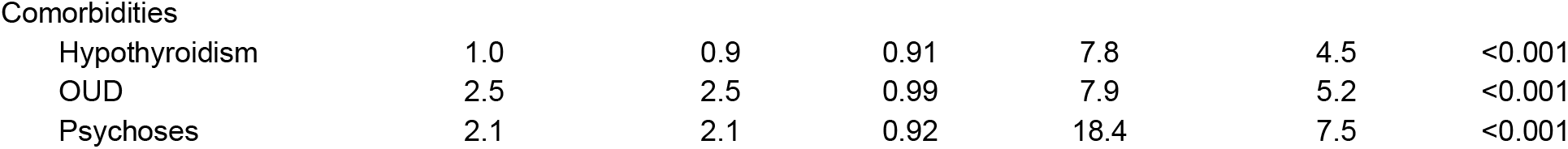
Demographic and clinical characteristics of patients with gunshot and stab recidivism. GSW-R, Gunshot wound recidivism. Stab-R, Stab wound recidivism OUD, opioid use disorder. TMPM, Trauma Mortality Prediction Model. TMPM score indicates predicted probability of death based on patient’s five most severe injuries.

**Figure 2.**
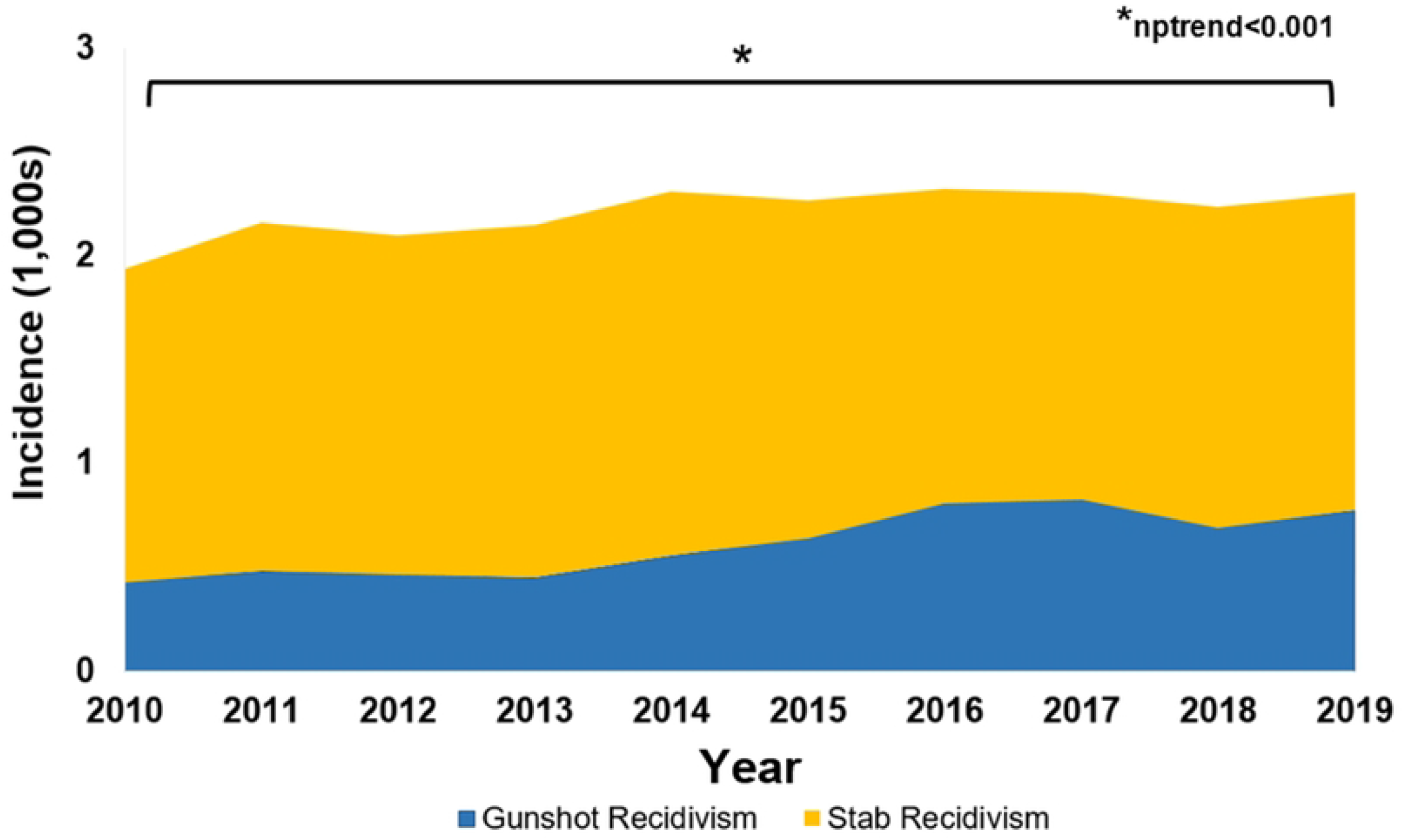
National trends in penetrating injury recidivism from 2010 to 2019.

**Figure 3.**
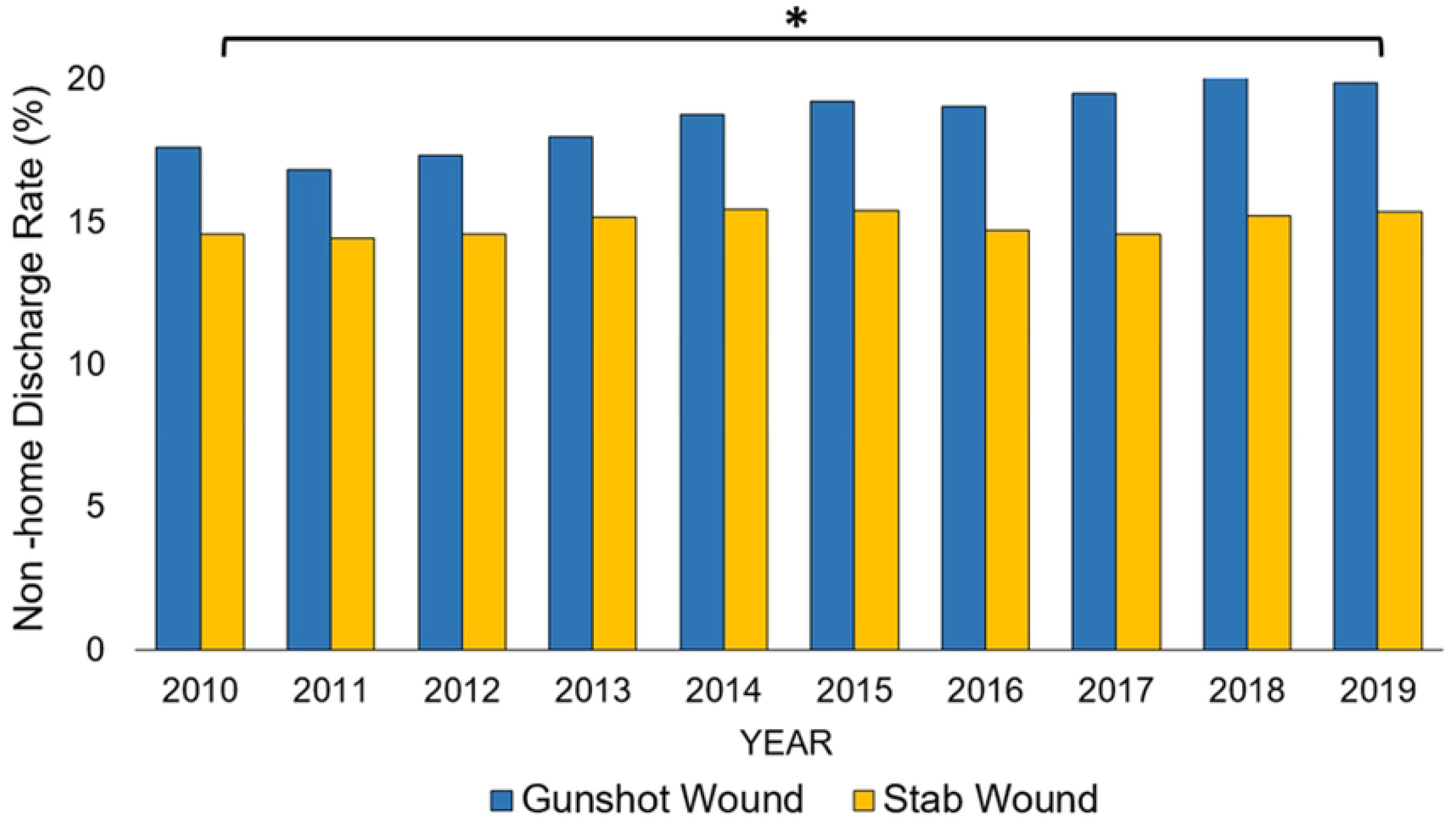
National trends in non-home discharge for patients experiencing penetrating trauma recidivism from 2010 to 2019.

### 3.2 Outcomes Associated with GSW Recidivism

Patients with GSW recidivism had shorter unadjusted index LOS (5 [2-10] vs 8 [4-18] days, P<0.001) and lower median index hospitalization cost ($18,000 [8,800-37,600] vs $28,800 [14,500-61,100], P<0.001), relative to others. Compared to those without recidivism, *GSW-R* had higher rates of non-home discharge (30.6 vs 25.9%, P<0.001). After risk adjustment, recidivism was associated with significantly higher cumulative hospitalization costs (β $19,200, 95% Confidence Interval [CI] $15,800-$22,300), a shorter estimated LOS by 0.7 days (95%CI 0.1-1.3 days), and increased odds of discharge to short-term care facility (Adjusted Odds Ratio [AOR] 5.4, 95%CI 4.3-6.8, Table 3). As shown in Figure 4, comorbidities including hypothyroidism, psychoses, and opioid use disorder were not associated with increased odds of gunshot injury recidivism.

**Figure 4.**
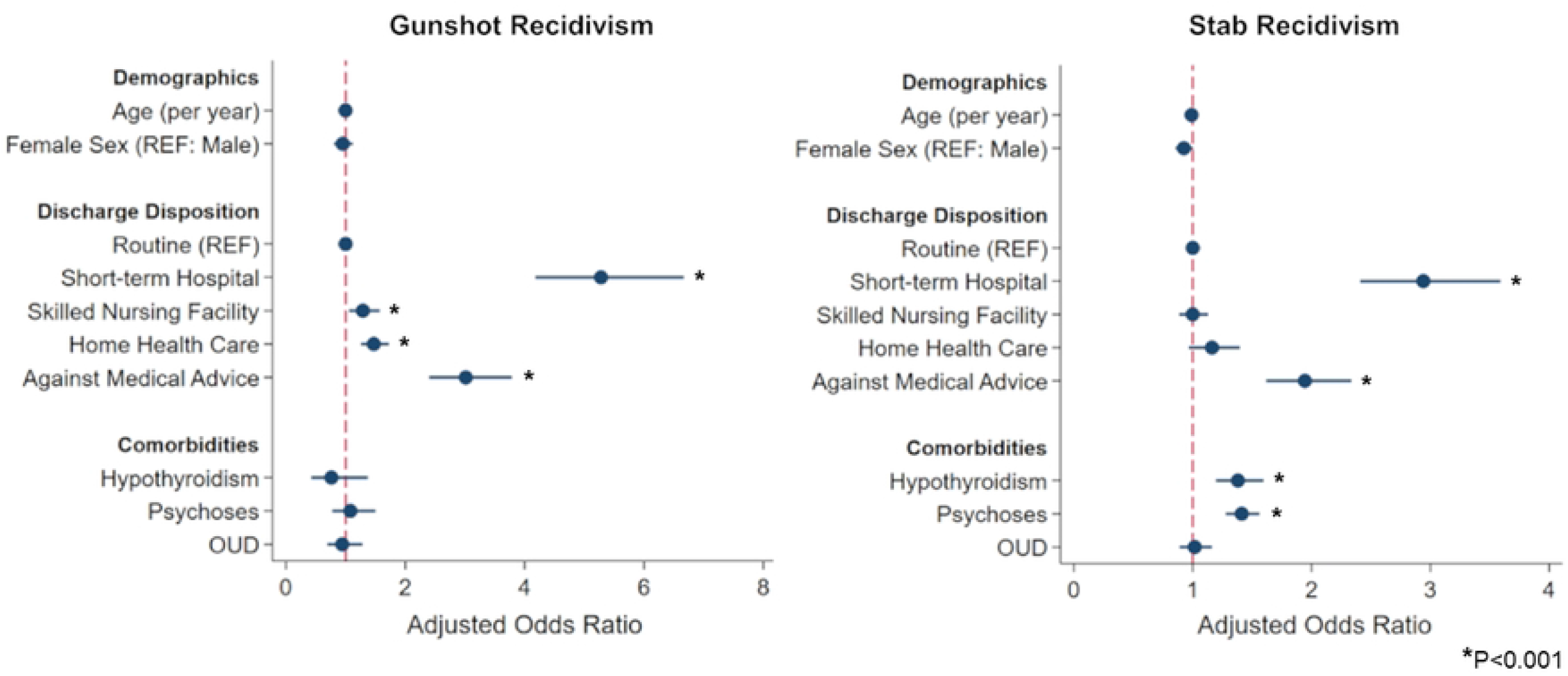
Risk-adjusted predictors of gunshot and stab injury recidivism. REF; reference. OUD, Opioid Use Disorder. Risk-adjusted predictors of stab injury recidivism. REF; reference. OUD, Opioid Use Disorder.

### 3.3 Demographics of Patients and Trends of Stab Recidivism

Of an estimated 687,863 patients, 15,142 (2.2%) was identified as *Stab-R* cohort. Over the study period, there was a stable trend in stab wound recidivism (nptrend=0.60, Figure 2). Similarly, non-home disposition among stab wound patients increased over the study period from 14.4% to 15.4% (nptrend <0.001, Figure3). Compared to those without repeated injury, patients from *Stab-R* were younger (36 [27-49] vs 44 [30-57] years, P<0.001), more often had an injury due to self-harm (65.8 vs 50.7%, P<0.001) and shorter median time until readmission (17 [5-54] vs 36 [11-95] days, P<0.001) compared to others. Furthermore, patients with recidivism had higher rates of depression (47.1 vs 30.1%, P<0.001), alcohol use disorder (24.7 vs 19.6%, P<0.001), and psychoses (18.4 vs 7.5%, P<0.001) relative to their non-recidivist counterparts (Table 1).

### 3.4 Outcomes Associated with Stab Recidivism

As shown in Table 2, underinsured and low-income status were associated with *Stab-R*. Those from *Stab-R* had shorter unadjusted index LOS (4 [2-7] vs 5 [3-8] days, P<0.001) and lower median index hospitalization cost ($6,600 [3,600-12,800] vs $7,900 [4,200-15,900], P<0.001) compared to others. Additionally, those with stab recidivism had higher rates of non-home discharge at index hospitalization (21.8 vs 17.3%, P<0.001) compared to others. After risk adjustment, *Stab-R* was associated with significantly higher cumulative hospitalization costs (β $13,300, 95% Confidence Interval [CI] $11,600-$15,000), shorter estimated LOS by 0.4 days (95%CI 0.4-0.6 days), and increased odds of transfer to short-term care facility (AOR 2.9, 95%CI 2.4-3.6) compared to home disposition (Table 3). Comorbid conditions of hypothyroidism (AOR 1.36, 95%CI 1.18-1.58) and psychoses (AOR 1.41, 95%CI 1.28-1.56) were associated with greater likelihood of *Stab-R* (Figure 4).

**Table 2.**
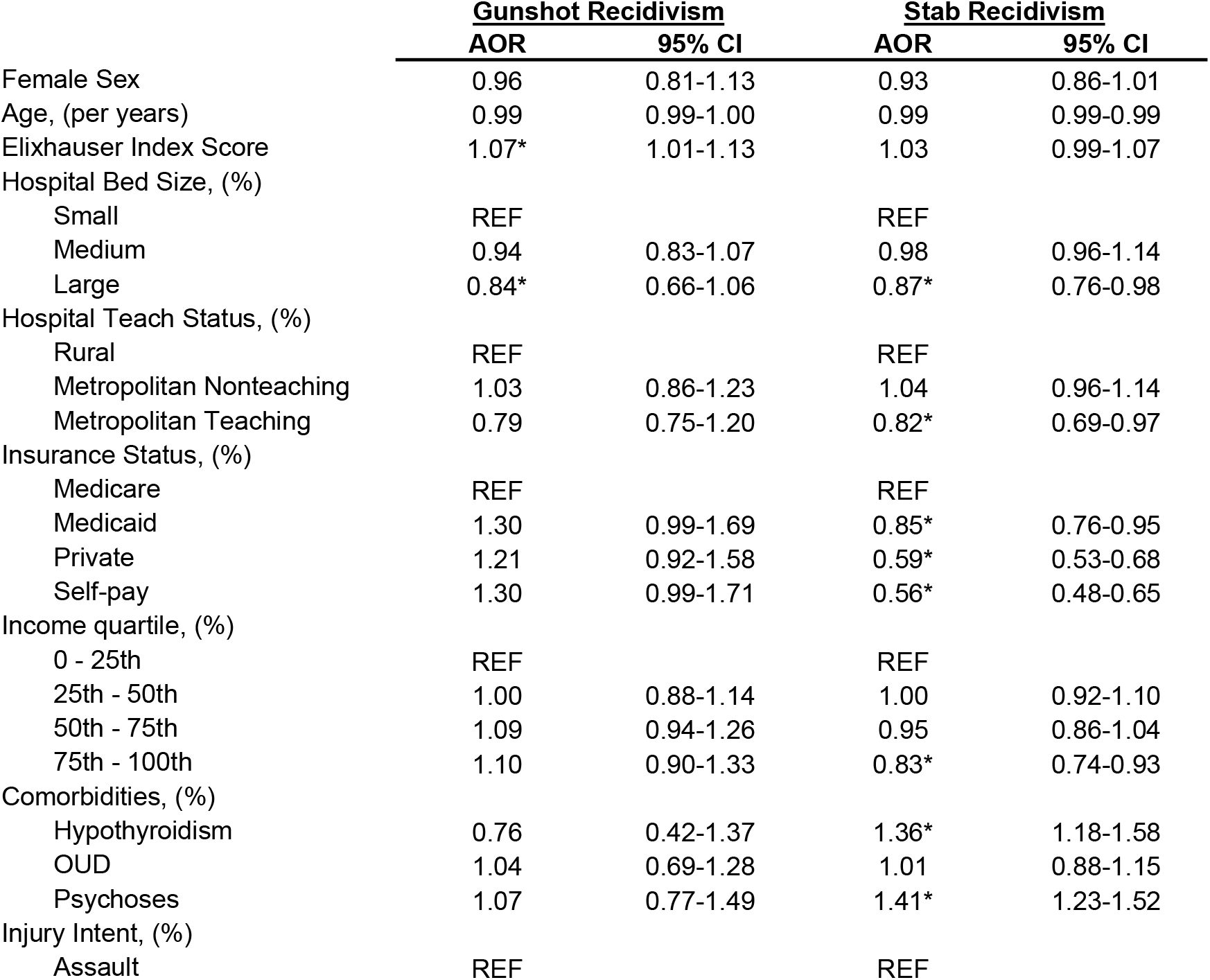
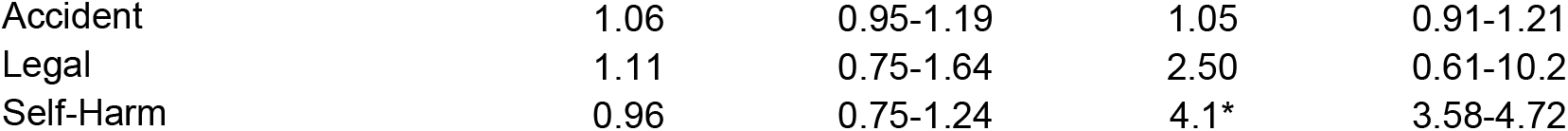
Risk-adjusted outcomes in patients with penetrating trauma recidivism. GSW-R, gunshot recidivism. Stab-R, stab recidivism. AOR, Adjusted Odds Ratio. CI, Confidence Interval. REF, Reference. OUD, Opioid Use Disorder.

|  | <u>Gunshot Recidivism</u> |  | <u>Stab Recidivism</u> |  |
| --- | --- | --- | --- | --- |
|  | <u>AOR</u> | <u>95% CI</u> | <u>AOR</u> | <u>95% CI</u> |
| Female Sex | 0.96 | 0.81-1.13 | 0.93 | 0.86-1.01 |
| Age, (per years) | 0.99 | 0.99-1.00 | 0.99 | 0.99-0.99 |
| Elixhauser Index Score | 1.07* | 1.01-1.13 | 1.03 | 0.99-1.07 |
| Hospital Bed Size, (%) |  |  |  |  |
| Small | REF |  | REF |  |
| Medium | 0.94 | 0.83-1.07 | 0.98 | 0.96-1.14 |
| Large | 0.84* | 0.66-1.06 | 0.87* | 0.76-0.98 |
| Hospital Teach Status, (%) |  |  |  |  |
| Rural | REF |  | REF |  |
| Metropolitan Nonteaching | 1.03 | 0.86-1.23 | 1.04 | 0.96-1.14 |
| Metropolitan Teaching | 0.79 | 0.75-1.20 | 0.82* | 0.69-0.97 |
| Insurance Status, (%) |  |  |  |  |
| Medicare | REF |  | REF |  |
| Medicaid | 1.30 | 0.99-1.69 | 0.85* | 0.76-0.95 |
| Private | 1.21 | 0.92-1.58 | 0.59* | 0.53-0.68 |
| Self-pay | 1.30 | 0.99-1.71 | 0.56* | 0.48-0.65 |
| Income quartile, (%) |  |  |  |  |
| 0 - 25th | REF |  | REF |  |
| 25th - 50th | 1.00 | 0.88-1.14 | 1.00 | 0.92-1.10 |
| 50th - 75th | 1.09 | 0.94-1.26 | 0.95 | 0.86-1.04 |
| 75th - 100th | 1.10 | 0.90-1.33 | 0.83* | 0.74-0.93 |
| Comorbidities, (%) |  |  |  |  |
| Hypothyroidism | 0.76 | 0.42-1.37 | 1.36* | 1.18-1.58 |
| OUD | 1.04 | 0.69-1.28 | 1.01 | 0.88-1.15 |
| Psychoses | 1.07 | 0.77-1.49 | 1.41* | 1.23-1.52 |
| Injury Intent, (%) |  |  |  |  |
| Assault | REF |  | REF |  |
| Accident | 1.06 | 0.95-1.19 | 1.05 | 0.91-1.21 |
| Legal | 1.11 | 0.75-1.64 | 2.50 | 0.61-10.2 |
| Self-Harm | 0.96 | 0.75-1.24 | 4.1* | 3.58-4.72 |

**Table 3.** Risk-adjusted outcomes in patients with gunshot and stab recidivism. Estimates are reported as AORs or β-coefficients for binary and continuous variables, respectively. CI; Confidence Interval; d; days.

|  | <u>Gunshot Recidivism</u> |  | <u>Stab Recidivism</u> |  |
| --- | --- | --- | --- | --- |
|  | Estimates | 95% CI | Estimates | 95% CI |
| LOS, (d) | -0.7 | -1.3--0.1 | -0.4 | -0.6--0.4 |
| Cumulative Cost, (\$1,000) | 19.1 | 15.8-22.3 | 13.3 | 11.6-15.0 |
| Discharge Location |  |  |  |  |
| Home | Reference |  | Reference |  |
| Short-Term Care Facility | 5.4 | 4.3-6.8 | 2.9 | 2.4-3.6 |
| Skilled Nursing Facility | 1.3 | 1.0-1.6 | 1.0 | 0.9-1.1 |
| Home Health | 1.5 | 1.3-1.7 | 1.2 | 0.9-1.4 |
| Against Medical Advise | 3.0 | 2.4-3.8 | 1.9 | 1.6-2.3 |

## 4. DISCUSSION

Penetrating trauma is the strongest predictor of future traumatic injury and is responsible for >$25 billion in annual costs based on medical expenses and lost productivity.^8^ Furthermore, the recurrence of gunshot and stab injuries underscores the prevalence of high-risk factors in a subset of the population at the highest risk for traumatic injuries in the United States. Previous studies have shown that penetrating trauma recidivism is likely not a random event but instead an indicator of elevated risk for excessive violence-related behaviors and exposures in the population.^1,3^ Importantly, patients presenting with recurrent penetrating trauma have significantly prolonged complications resulting from the injury and have a 77% higher likelihood of long-term mortality following recidivism.^2^ Nonetheless, data regarding trends, LOS, hospitalization expenditure, non-home discharge rate and factors associated with penetrating trauma recidivism is lacking in the literature. In the present work, we noted an increasing national trend in gunshot recidivism with nearly 2.1% experiencing this event within 60 days of initial injury. Over the study period, the incidence of stab recidivism remained similar. We also found patients with non-home disposition to face increased odds of re-injury. Several of these findings warrant further discussion.

Consistent with a rise in firearm-related morbidity and mortality in the US, we noted a significant escalation of gunshot injury recidivism over the study period. Non-fatal GSWs are two-fold more common than fatal injuries, and surviving victims are predisposed to a cycle of violence.^9^ Among patients experiencing gunshot recidivism, present study showed that 91.8% of initial gunshot wounds occur due to assault or accident. These findings highlight the prevalence of urban violence resulting from inadequate firearm regulation. Policies to promote gun safety and laws that mitigate gun-related danger in communities are necessitated to prevent recurrent gun injury in high-risk patients. Moreover, the lack of gun-related injury prevention efforts among perioperative physicians has also been proposed as an area of improvement.^10,11^ Healthcare professionals are frequently the first or only caregivers to come in contact with victims of firearm-related injury. As such, it is essential for physicians to identify patients at risk for recidivism. Holistic interventions involving physicians controlling substance use, providing psychiatric care for mood disorders and facilitating prevention measures may help reduce gunshot recidivism.

In contrast to gunshot injury recidivism, the respective rates for stab injury have remained steady over the past decade. Interestingly, we found a significant association between stab recidivism and comorbid conditions such as psychoses and hypothyroidism. We further noted that 68.8% of patients experiencing stab recidivism were injured due to self-harm. Self-injurious behaviors are strongly associated with mental disorders, including psychosis and depression.^12,13^ Borde and colleagues have shown increased odds of clinical depression with any severity of hypothyroidism.^14^ In addition to clinical depression, dysregulation of thyroid function can manifest with a wide range of psychiatric symptoms, including altered personality with psychotic symptoms, both of which can ultimately lead to self-harming behaviors.^15^ These findings highlight potentially modifiable factors that increase the risk for recurrent stab injuries. It is also important to highlight low socioeconomic status and a prolonged period of untreated psychiatric illness as contributory to risk of self-harm.^16^ In the present study, we noted Medicare or Medicaid coverage and low income status to be a risk factor for stab wound recidivism. Although the exact nature of the association remains unclear, it is not unreasonable to assume that underserved patients lack longitudinal care that may mitigate high-risk behaviors or urban trauma exposure. Easier provider access to screening tools or adoption of a trauma-informed approach may reduce recidivism in patients with concomitant psychiatric illnesses or inadequate access to resources.

Regardless of the primary penetrating trauma, we found non-home discharge to be associated with higher rates of recidivism. Leaving against medical advice can limit appropriate rehabilitation, psychiatric evaluation and social services in patients with a traumatic injury. Ibrahim et al. showed younger age, Black race, low household income, and lack of medical insurance as predictors of being discharged against medical advice.^17^ These factors align with risk factors for trauma and a lower likelihood of receiving follow-up care and rehabilitation.^18^ Prior direct or indirect violence exposure is highly prevalent among patients experiencing recidivism, often intensifying the post-traumatic stress symptoms of sleep disturbance and depression.^19^ Being transferred to a short-term medical facility restrains a proper psychiatric evaluation and biopsychosocial treatment in patients, which may be necessary to prevent self-harming behaviors. Treatment for penetrating injuries at trauma centers having expertise in the management of such patients and scheduling adequate follow-up care, may reduce this recidivism.^20,21^ Nevertheless, substantial geographical locations and variations in emergency medical service availability often hinder access to trauma center services, leaving patients with limited options for discharge.^22^ Taken into the context of risk for recidivism, institutional programs to provide longitudinal post-trauma monitoring and treatment may benefit patients following penetrating trauma. In addition, intervention and prevention programs targeting gun violence in urban and rural areas are warranted to reduce recurrent injuries among trauma patients.

This study had several important limitations due to its retrospective design and nature of the NRD. Although the AHRQ has quality control measures to ensure best practices for coding, the administrative nature of the NRD allows the potential for miscoded events and missing observations within the database. Additionally, the NRD lacks surgeon-specific variables as well as patient-identifying variables, such as race, location and hospital data. The NRD also lacks granular details of readmission between states, medication use, and the presence of a pain team consult. Although the strength of the database to track repeat hospitalizations at participating hospitals, long-term survival and out-of-hospital mortality are unable to be determined. We were unable to delineate affiliations between care facilities and reasons for non-home discharge dispositions. Lastly, hospital-level analysis was limited as we were unable to evaluate trauma center designation. Despite these limitations, we utilized statistically robust methodologies to reduce bias and evaluated penetrating trauma recidivism using a nationally representative cohort. Our findings highlight multiple factors that may supplement the prevention interventions in high-risk patients of penetrating trauma recidivism.

The rate of recidivism among patients with penetrating trauma has increased over the past decade and continues to pose clinical and financial burden in the US. Without deliberate action, the cyclic pattern of re-injury will not abate but solely pose a long-term risk of morbidity and mortality in these patients. Therefore, national efforts to improve post-discharge care and social support services for patients with penetrating trauma are warranted. Furthermore, multidisciplinary approaches involving physicians, public health leaders and legislators are required to reduce the national burden of recidivism.

## Data Availability

All relevant data are within the manuscript and its Supporting Information files.

## Study Type

Retrospective cohort study

## Author Contributions

**Nam Yong Cho**: Conceptualization, Methodology, Data Analysis, Writing Original Draft. **Russyan Mabeza**: Validation, Writing, Reviewing, Editing. **Syed Shahyan Bakhtiyar:** Conceptualization, Validation, Editing. **Shannon Richardson**: Writing, Reviewing, Editing. **Konmal Ali:** Writing, Generating Tables/Figures. **Zachary Tran:** Conceptualization, Validation, Reviewing. **Peyman Benharash:** Conceptualization, Methodology, Supervision, Reviewing

## Availability of Data

The data that support the findings of this study are available from the Healthcare Cost and Utilization Project (HCUP). Restrictions apply to the availability of these data, which were used with permission for this study. Data are available from the authors with the permission of the Healthcare Cost and Utilization Project.

## MEETING PRESENTATION

Accepted for ***Oral Presentation*** at the 18^th^ Annual Academic Surgical Congress.

Abstract #: ASC20230411

Session: 37 – Clinical/Outcomes: Trauma/Critical Care Oral Session I

Date/Location: Wednesday, February 8, 2023 / Houston, Texas

## FIGURE LEGENDS

**Supplemental Table S1.** International Classification of Disease, Ninth and Tenth revision, Codes (ICD-9/10) for Penetrating Traumas.

| Characteristics | ICD-9-CM Codes | ICD-10-CM codes |
| --- | --- | --- |
| Gunshot Injury | E965x, E922x,<br>E955x, E970,E9794,<br>E985x | X940XXA, X941XXA, X942XXA, X948XXA, X949XXA,<br>X9501XA, X9502XA, X9509XA, X958XXA, X959XXA,<br>X93XXXA, W320XXA, W321XXA, W3301XA, W3300XA,<br>W3302XA, W3303XA, W3309XA, W3310XA, W3311XA,<br>W3312XA, W3313XA, W3319XA, W3400XA, W34010A,<br>W34011A, W34018A, W3409XA, W3410XA, W34110A,<br>W34111A, W34118A, W3419XA, X72XXXA, X730XXA,<br>X731XXA, X732XXA, X739XXA, X738XXA, X7401XA,<br>X7402XA, X7409XA, X748XXA, X749XXA, Y35003A,<br>Y35002A, Y35009A, Y35011A, Y35012A, Y35013A,<br>Y35019A, Y35021A, Y35022A, Y35023A, Y35029A,<br>Y35031A, Y35032A, Y35033A, Y35039A, Y35041A,<br>Y35042A, Y35043A, Y35049A, Y35091A, Y35092A,<br>Y35093A, Y35099A, Y22XXXA, Y230XXA, Y231XXA,<br>Y232XXA, Y233XXA, Y238XXA, Y239XXA, Y240XXA,<br>Y248XXA, Y249XXA |
| Stab Injury | E919x, E966, E974,<br>E956, E920x, E919x | X990XXA, X991XXA, X992XXA, X998XXA, X999XXA,<br>Y35401A, Y35402A, Y35403A, Y35409A, Y35411A,<br>Y35412A, Y35413A, Y35419A, Y35491A, Y35492A,<br>Y35493A, Y35499A, W240XXA, W241XXA, W25XXXA,<br>W260XXA, W261XXA, W262XXA, W268XXA, W269XXA,<br>W28XXXA, W293XXA, W292XXA, W294XXA, W291XXA,<br>W298XXA, W300XXA, W301XXA, W302XXA, W303XXA,<br>W3081XA, W3089XA, W309XXA, W310XXA, W311XXA,<br>W312XXA, W313XXA, W3181XA, W3182XA, W3183XA,<br>W3189XA, W319XXA, X780XXA, X781XXA, X782XXA,<br>X788XXA, X789XXA |

**Supplemental Table S2.**
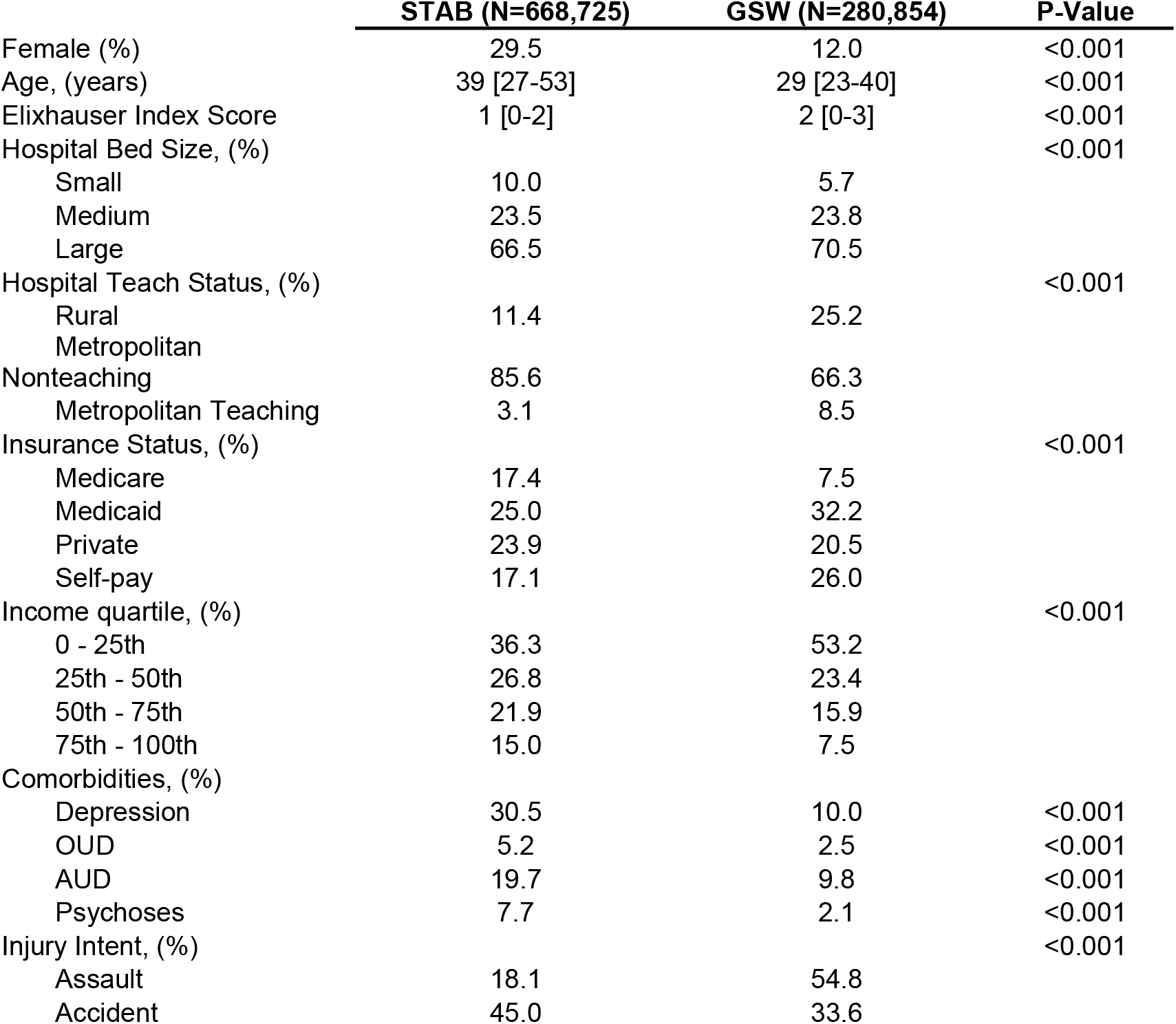
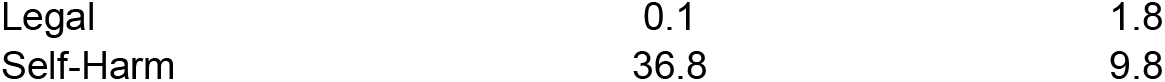
Demographic and Clinical Characteristics of Patient with Penetrating Injury Stratified by Mechanism of Injury

|  | **STAB (N=668,725)** | **GSW (N=280,854)** | **P-Value** |
| --- | --- | --- | --- |
| Female (%) | 29.5 | 12.0 | <0.001 |
| Age, (years) | 39 [27-53] | 29 [23-40] | <0.001 |
| Elixhauser Index Score | 1 [0-2] | 2 [0-3] | <0.001 |
| Hospital Bed Size, (%) |  |  | <0.001 |
| Small | 10.0 | 5.7 |  |
| Medium | 23.5 | 23.8 |  |
| Large | 66.5 | 70.5 |  |
| Hospital Teach Status, (%) |  |  | <0.001 |
| Rural | 11.4 | 25.2 |  |
| Metropolitan |  |  |  |
| Nonteaching | 85.6 | 66.3 |  |
| Metropolitan Teaching | 3.1 | 8.5 |  |
| Insurance Status, (%) |  |  | <0.001 |
| Medicare | 17.4 | 7.5 |  |
| Medicaid | 25.0 | 32.2 |  |
| Private | 23.9 | 20.5 |  |
| Self-pay | 17.1 | 26.0 |  |
| Income quartile, (%) |  |  | <0.001 |
| 0 - 25th | 36.3 | 53.2 |  |
| 25th - 50th | 26.8 | 23.4 |  |
| 50th - 75th | 21.9 | 15.9 |  |
| 75th - 100th | 15.0 | 7.5 |  |
| Comorbidities, (%) |  |  |  |
| Depression | 30.5 | 10.0 | <0.001 |
| OUD | 5.2 | 2.5 | <0.001 |
| AUD | 19.7 | 9.8 | <0.001 |
| Psychoses | 7.7 | 2.1 | <0.001 |
| Injury Intent, (%) |  |  | <0.001 |
| Assault | 18.1 | 54.8 |  |
| Accident | 45.0 | 33.6 |  |

|  |  |  |
| --- | --- | --- |
| Legal | 0.1 | 1.8 |
| Self-Harm | 36.8 | 9.8 |

**Supplemental Figure 1.**
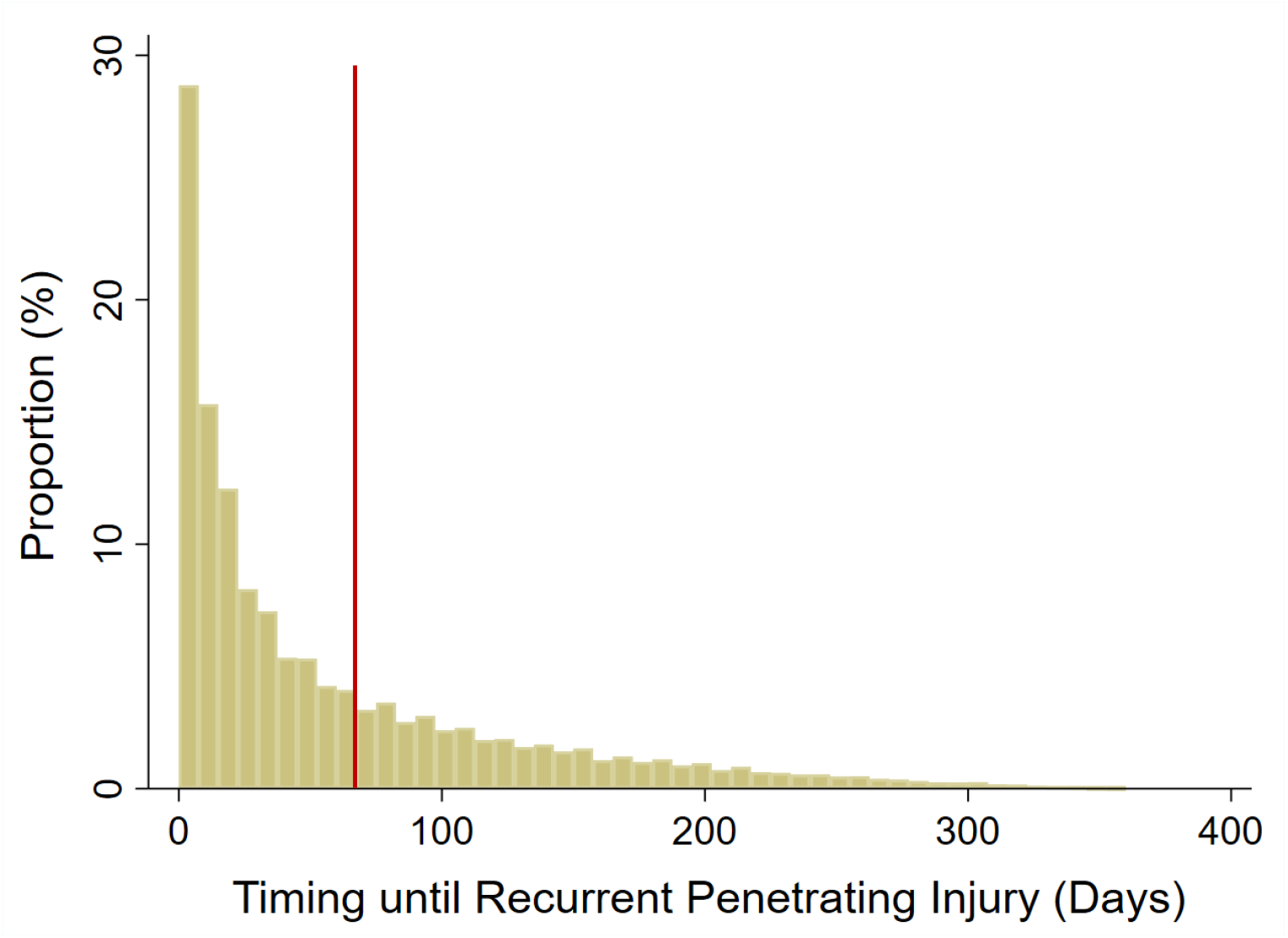
Proportion of timing until recurrent penetrating injury in patients with recidivism (Referenece line set at 60 Days). TO BE UPDATED.

## Reference

1. Kao AM, Schlosser KA, Arnold MR, et al. Trauma recidivism and mortality following violent injuries in young adults. J Surg Res. 2019;237:140–147. doi:10.1016/j.jss.2018.09.006

2. Strong BL, Greene CR, Smith GS. Trauma recidivism predicts long-term mortality: missed opportunities for prevention(Retrospective cohort study). Ann Surg. 2017;265(5):847–853. doi:10.1097/SLA.0000000000001823

3. Brooke BS, Efron DT, Chang DC, Haut ER, Cornwell EE. Patterns and outcomes among penetrating trauma recidivists: it only gets worse. J Trauma. 2006;61(1):16–19; discussion 20. doi:10.1097/01.ta.0000224143.15498.bb

4. Osler TM, Glance LG, Cook A, Buzas JS, Hosmer DW. A trauma mortality prediction model based on the ICD-10-CM lexicon: TMPM-ICD10. J Trauma Acute Care Surg. 2019;86(5):891–895. doi:10.1097/TA.0000000000002194

5. Nrd description of data elements. Accessed December 20, 2022. https://www.hcup-us.ahrq.gov/db/nation/nrd/nrddde.jsp

6. Elixhauser A, Steiner C, Harris DR, Coffey RM. Comorbidity measures for use with administrative data. Med Care. 1998;36(1):8–27. doi:10.1097/00005650-199801000-00004

7. Zou H, Hastie T. Regularization and variable selection via the elastic net. J Royal Statistical Soc B. 2005;67(2):301–320. doi:10.1111/j.1467-9868.2005.00503.x

8. Pino EC, Fontin F, James TL, Dugan E. Mechanism of penetrating injury mediates the risk of long-term adverse outcomes for survivors of violent trauma. J Trauma Acute Care Surg. 2022;92(3):511–519. doi:10.1097/TA.0000000000003364

9. Fontanarosa PB, Bibbins-Domingo K. The unrelenting epidemic of firearm violence. JAMA. 2022;328(12):1201. doi:10.1001/jama.2022.17293

10. McGraw C, Jarvis S, Carrick M, et al. Examining trends in gun violence injuries before and during the COVID-19 pandemic across six trauma centers. Trauma Surg Acute Care Open. 2022;7(1):e000801. doi:10.1136/tsaco-2021-000801

11. Gerstein NS, Sanders JC, McCunn M, et al. The gun violence epidemic: time for perioperative physicians to act. Journal of Cardiothoracic and Vascular Anesthesia. 2018;32(3):1097–1100. doi:10.1053/j.jvca.2018.03.002

12. Honings S, Drukker M, Groen R, van Os J. Psychotic experiences and risk of self-injurious behaviour in the general population: a systematic review and meta-analysis. Psychol Med. 2016;46(2):237–251. doi:10.1017/S0033291715001841

13. Hielscher E, DeVylder JE, Saha S, Connell M, Scott JG. Why are psychotic experiences associated with self-injurious thoughts and behaviours? A systematic review and critical appraisal of potential confounding and mediating factors. Psychol Med. 2018;48(9):1410–1426. doi:10.1017/S0033291717002677

14. Bode H, Ivens B, Bschor T, Schwarzer G, Henssler J, Baethge C. Association of hypothyroidism and clinical depression: a systematic review and meta-analysis. JAMA Psychiatry. 2021;78(12):1375. doi:10.1001/jamapsychiatry.2021.2506

15. Heiberg Brix T, Ferløv-Schwensen C, Thvilum M, Hegedüs L. Death by unnatural causes, mainly suicide, is increased in patients with Hashimoto’s thyroiditis. A nationwide Danish register study. Endocrine. 2019;65(3):616–622. doi:10.1007/s12020-019-01946-5

16. Harvey SB, Dean K, Morgan C, et al. Self-harm in first-episode psychosis. Br J Psychiatry. 2008;192(3):178–184. doi:10.1192/bjp.bp.107.037192

17. Ibrahim SA, Kwoh CK, Krishnan E. Factors associated with patients who leave acute-care hospitals against medical advice. Am J Public Health. 2007;97(12):2204–2208. doi:10.2105/AJPH.2006.100164

18. Haider AH, Weygandt PL, Bentley JM, et al. Disparities in trauma care and outcomes in the United States: A systematic review and meta-analysis. Journal of Trauma and Acute Care Surgery. 2013;74(5):1195–1205. doi:10.1097/TA.0b013e31828c331d

19. Corbin T, Tabb LP, Waite D, et al. Posttraumatic stress, depression, and sleep among young survivors of violence. Journal of Health Care for the Poor and Underserved. 2021;32(3):1339–1358. doi:10.1353/hpu.2021.0136

20. Gibson PD, Ippolito JA, Shaath MK, Campbell CL, Fox AD, Ahmed I. Pediatric gunshot wound recidivism: Identification of at-risk youth. Journal of Trauma and Acute Care Surgery. 2016;80(6):877–883. doi:10.1097/TA.0000000000001072

21. Jarman MP, Hashmi Z, Zerhouni Y, et al. Quantifying geographic barriers to trauma care: Urban-rural variation in prehospital mortality. J Trauma Acute Care Surg. 2019;87(1):173–180. doi:10.1097/TA.0000000000002335

